# A New Adaptive Logistic Model for Epidemics and the Resurgence of COVID-19 in the United States

**DOI:** 10.1101/2020.07.17.20156109

**Authors:** David H. Roberts

**Affiliations:** Department of Physics MS-057, Brandeis University, Waltham, MA 02454-0911 USA

## Abstract

The Adaptive Logistic Model (ALM) of epidemics incorporates the results of infection mitigation effects on the course of an epidemic, and well describes the histories of the COVID-19 epidemics in many countries, including the United States. In particular, it is much more successful than is a basic logistic model. However, in the U.S. these mitigation efforts have recently been relaxed in many places, resulting in the second peak in infections that started in late May of 2020. In this paper the ALM is modified to account for the relaxation of these mitigation effects, leading to the Adaptive Logistic Model 2 (ALM-2). The ALM-2 is then used to understand quantitatively the second peak of COVID-19 cases. The ALM-2 is also successfully applied to the data on deaths even though they do not yet show a second peak.

## 1. Introduction

The United States has been particularly impacted by the novel coronavirus (SARS-CoV-2) and the disease it causes, COVID-19. In the U.S. the daily counts of cases rose sharply starting the second week of March 2020 (approximately day 75), and reached a peak about 25 days later. It then began to decline slowly and did so for about 45 days. However, second peak began to grow starting in the fourth week of May (about day 145), and continues to grow at this time (July 15, 2020, day 197). See Fig. 1 for plots of the time histories of cases and deaths in the U.S.

**Fig. 1.**
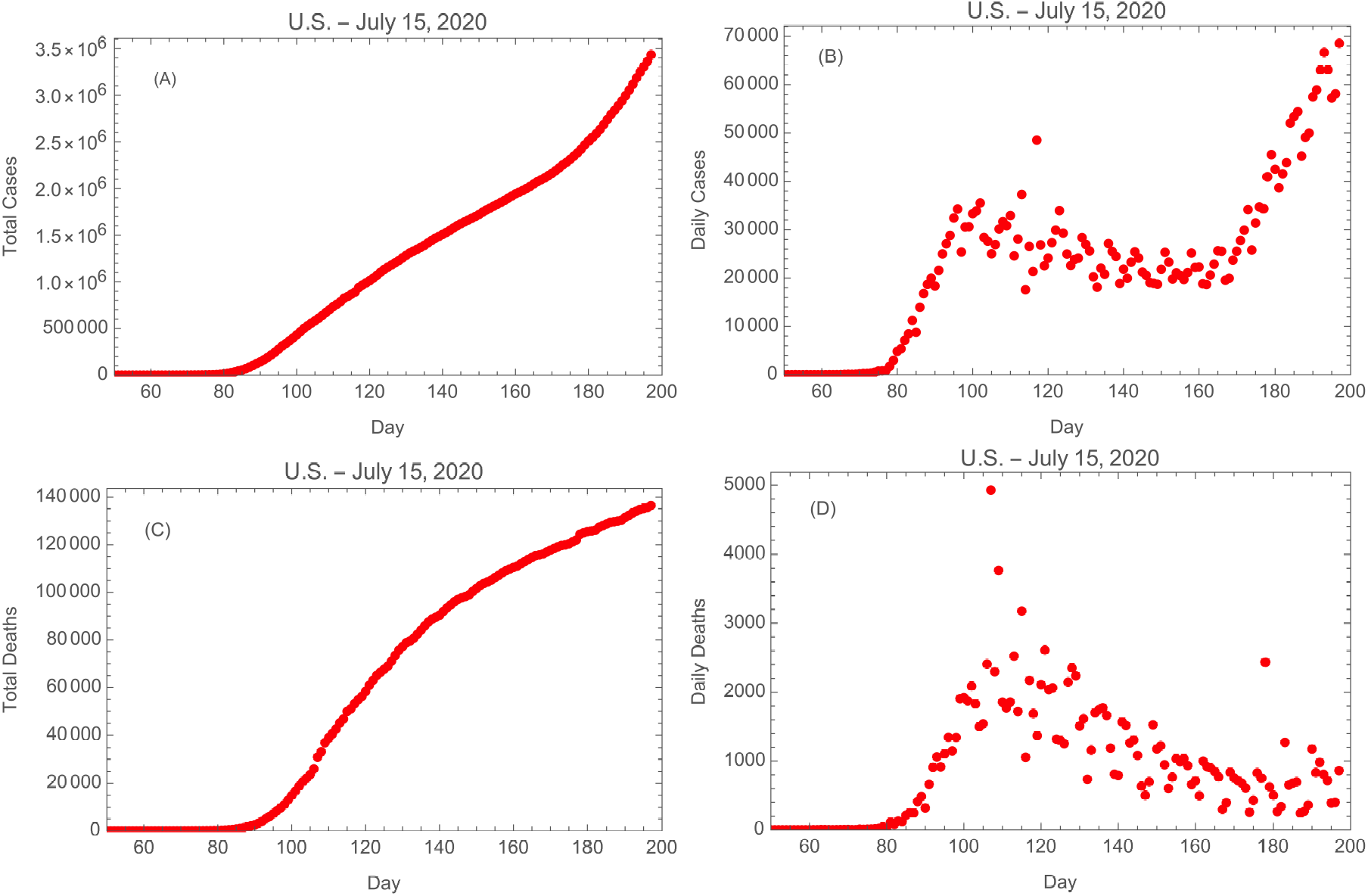
U.S. COVID-19 data as of July 15, 2020: (A) Total cases, (B) daily cases, (C) total deaths, & (D) daily deaths. Day 1 is January 1, 2020.

Roberts (2020a) demonstrated that the details of the growth and subsequent decline of the counts of daily cases could be understood using two new epidemiological models, the Distributed Logistic Model (DLM) and the Adaptive Logistic Model (ALM). In particular, the ALM ascribes the details of the growth and decline of the daily counts to mitigation efforts that reduced the chance that an infected person would infect others. The current paper presents an enhancement of the ALM, the Adaptive Logistic Model 2 (ALM-2), that incorporates the relaxation of these mitigation efforts, and shows that this accounts quantitatively for the details of the second peak of infections. For the details of the derivations and applications of the Basic Logistic Model, the DLM, and the ALM, the reader is referred to Roberts (2020a).

The plan of this paper is as follows. In Section 2 we review the motivation for the ALM. In Section 3 its modification, the ALM-2, is presented. It is shown to well-describe the details of the second peak of infection in the U.S. in Section 4. Section 5 discusses the fits to the deaths data, Section 6 summarizes the results, and Section 7 has the conclusions.

## 2. The Adaptive Logistic Model

### 2.1. Derivation

A classic model for epidemics is described by the *logistic differential equation* for the number *f* of infected individuals,

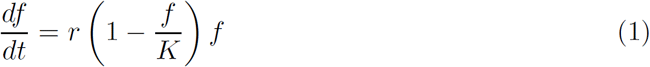

(see Hethcote 2000 for a review of the mathematics of epidemics). This represents exponential growth tempered by the effects of a finite population *K* of succeptable individuals. Here *r* is the rate constant. The solution of this equation is

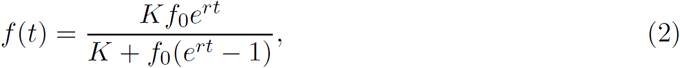

which satisfies the conditions *f* (0) = *f*_0_ and *f* (∞) = *K*. The time course described by Eq. 2 is the familiar “S curve” used to describe bacterial growth and other rate limited phenomena. The history of the total number of cases of disease *f* (*t*) can be fitted by this equation to determine the parameters *r* and *K*, each of which has a well-defined meaning. In particular, *K* represents a prediction of the endpoint of the epidemic. Unfortunately, as shown in Roberts (2020a), Eq. 2 does not provide a good description of the COVID-19 epidemic in the U.S. or other countries.

During the course of the COVID-19 pandemic, countries across the world introduced a number of measures to reduce the spread of the disease, including quarantining, masking, social distancing, and contact tracing and isolating. As the pandemic progressed these measures tended to become more effective. Their effect was to make the average infected individual progressively less likely to infect another person. A factor that reduces the probability of transmission as the pandemic progresses can be introduced into Eq. 1 to account for this effect. A simple way to do this is to modify the rate constant *r* by a function that decreases as the infected population increases. Following Roberts (2020a), the function *S/*(*S* + *g*) = 1/(1 + *g/S*), where *S* is a dimensionless parameter to be determined from the data, is used because it yields an excellent fit to the data. This leads to the *adaptive logistic equation*^2^

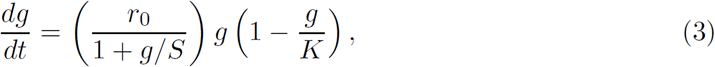

where now *g*(*t*) is the number of infected individuals. Here *r*_0_ is the rate constant at the beginning of the epidemic (*g* ≪ *S*). When *g* reaches *S* the initial rate constant is halved, so in that sense *S* is the half-life of the effective rate constant. The function

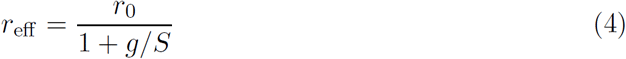

is the effective rate constant for this model; as the population *g* increases *r*_eff_ decreases, so r_eff_ is a function of time.

### 2.2. Method of Solution

Solution of Eq. 3 yields a function *g*(*t*) that depends on the four parameters (*g*_0_, *r, K, S*), where *g*_0_ is the initial value *g*(0). By minimizing the summed squares of the differences between the function *g*(*t*) and the total counts of cases or deaths one can find estimates for these parameters. A closed form solution to Eq. 3 could not be found, so it was solved and fit to the data numerically. This was done with Mathematica™ code as shown in Roberts (2020a). This is time consuming because it involves repeated numerical solutions of the differential equation (3). To determine the predicted distributions of the number of daily cases or deaths one takes the numerical derivatives of the fits to *g*(*t*). This does not fit the daily numbers, it merely compares them with the expectations derived from the fits to the total numbers. The fits of this model to the cases and deaths data in the U.S., Italy, and the United Kingdom are shown in Roberts (2020a).

## 3. Adaptive Logistic Model 2

In late May 2020 the number of cases of COVID-19 in the U.S. began to deviate from its gradual decline of two months, and started to peak sharply (see Fig. 1B). One interpretation of this event is that the moderating effects of social distancing, etc., are beginning to wane, and as a result the effective rate constant is beginning to rise. To model this a mathematical representation of an effective rate constant that initially declines and then starts to climb was sought. Some experimentation led to the form^3^

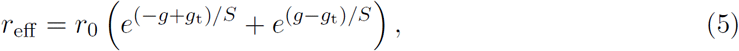

where *g* is the number of cases and (*g*_t_, *S*) are parameters to be determined from the data. Here *g*_t_ represents the value of *g* where the transition from falling to rising effective rate constant occurs, and *S* determines its rate of change. Using the parameters determined below from the cases and deaths data, *r*_eff_ (*g*) is illustrated in Fig. 2.

**Fig. 2.**
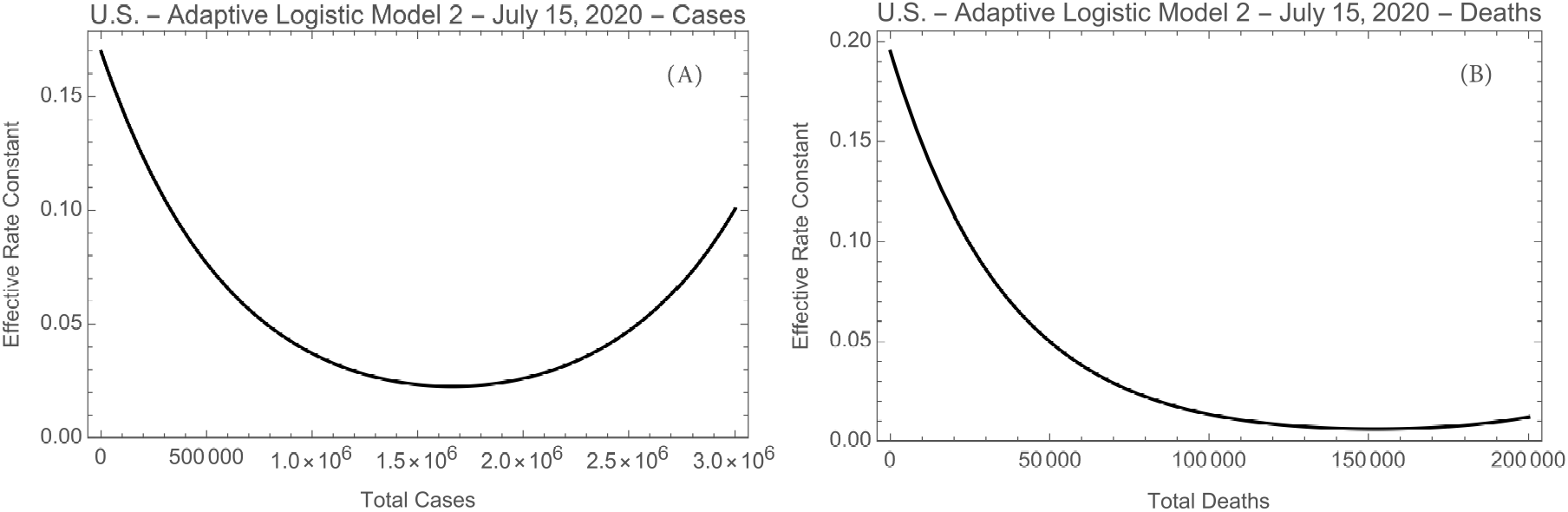
The effective rate constant as a function of the number of (A) total number of cases, and (B) total number of deaths.

With this change the ALM becomes the *Adaptive Logistic Model 2* (ALM-2), governed by the non-linear differential equation

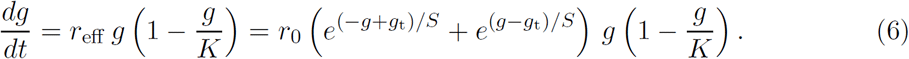

## 4. Application to Cases in the U.S., July 2020

### 4.1. Method of Solution 1

Now Eq. 6 is applied to the most recent U.S. data, taken from ECDPC (2020); they span January 1, 2020 through July 15, 2020 (197 days). Rather than attempt to determine the parameters of the model by repeated numerical solutions of this differential equation using only the total cases data, as described in Section 2.2, instead one can combine the daily and total data by minimizing the sum of the squares of the differences between the left hand side and the right hand side of Eq. 6 for each day, using the fact that *dg/dt* represents the daily numbers. Let *n*_i_ be the *N*_data_ = 197 daily cases numbers and *N*_i_ be the *N*_data_ total cases numbers in the data set; then the quantity to be minimized is

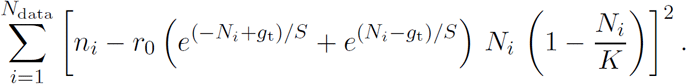

This must be minimized simultaneously with respect to the four parameters (*r*_0_, *K, S, g*_t_); this was done with the Mathematica™ function NMinimize.

The final result of this exercise is found by inserting the best fit parameters into the right hand side of Eq. 6, thus giving values for the left hand side, that is, for the daily numbers of cases (the fitted values of *dg/dt* for each day). The modeled total numbers are obtained by summing the modeled daily numbers. Figs. 3 shows comparison of the data with the model fits for the total and daily numbers. The main features to note are that the second peak in the daily data is well reproduced in the model, as is the corresponding kink in the total cases curve. The best fit parameters are presented in Table 1.

**Fig. 3.**
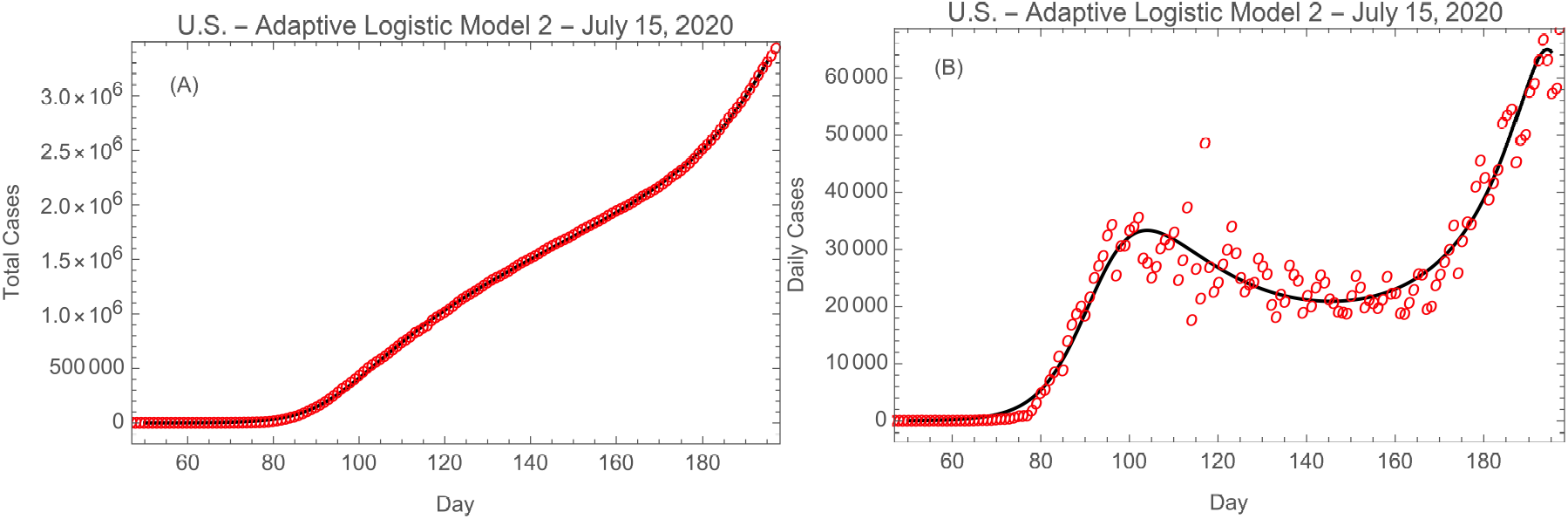
ALM-2 fit to (A) total cases and (B) daily cases of COVID-19 in the U.S. The red circles are the data and the solid black lines are the model fits. Day 1 is January 1, 2020.

**Table 1.** Parameters of the Adaptive Logistic Model 2

| Parameter | Units | Cases | Deaths |
| --- | --- | --- | --- |
| $N_{\text{data points}}$ | ... | 197 | 197 |
| $N_{\text{tot}}$ | ... | 3,304,942 | 135,205 |
| $K$ | ... | $3.89 \times 10^6$ | $1.08 \times 10^7$<sup>a</sup> |
| $r_0$ | day<sup>-1</sup> | 0.00110 | 0.00180 |
| $r_{\text{eff}}(g = 0)$ | day<sup>-1</sup> | 0.170 | 0.195 |
| $S$ | ... | $6.25 \times 10^5$ | $3.33 \times 10^4$ |
| $g_t$ | ... | $1.69 \times 10^6$ | $1.57 \times 10^5$ |

<sup>a</sup>This value is poorly constrained.

### 4.2. Method of Solution 2

As a check, the solution to Eq. 6 was also done as described in Section 2.2 above, using only the total cases data. The results were essentially indistinguishable from those found in Section 4.1 above; in each case we use the results from method 2 because it yields functional forms for the solutions (as numerical interpolation functions).

## 5. Application to Deaths in the U.S., July 2020

The same methods described above can be applied to the history of deaths in the U.S. (data also taken from ECDPC 2020). The results are shown in Fig. 4, and the best fit parameters are presented in Table 1.

**Fig. 4.**
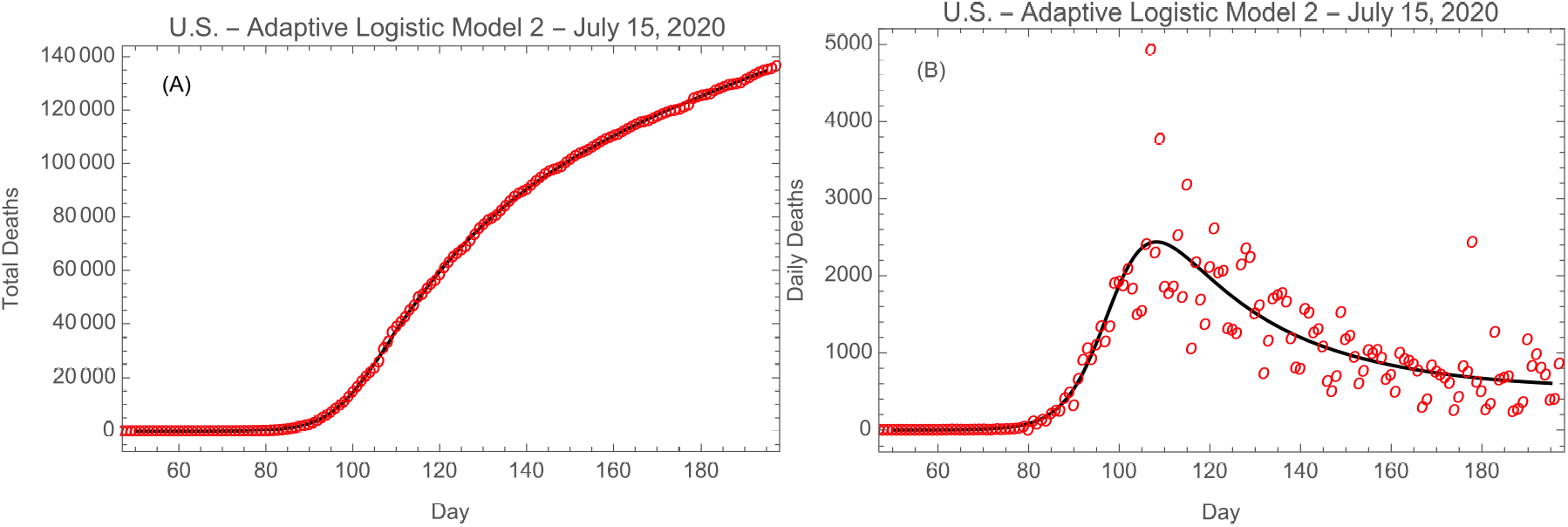
ALM-2 fit to (A) total deaths and (B) daily deaths due to COVID-19 in the U.S. The red circles are the data and the solid black lines are the model fits. Day 1 in January 1, 2020.

The ALM-2 fits to daily cases and deaths are shown together in Fig. 5. Comparing the parts of these curves between days 60 and 120, the delay between the rises of the two curves is 6 ± 1 days, and the mean mortality ratio is 7.1 ± 0.1% ≃ 1/14. This is illustrated in Fig. 6B. If instead we compare the total cases and deaths curves, the shift is 8 ± 1 days and the mean mortality ratio is 7.6 ± 0.4% ≃ 1/13; see Fig. 6A. These numbers represent the mortality ratios during the initial growing phase of the epidemic. As can be inferred from the subsequent divergence of the two curves in Fig. 5, the mortality ratio has fallen significantly since the time of the first peak (see also Roberts 2020a).

**Fig. 5.**
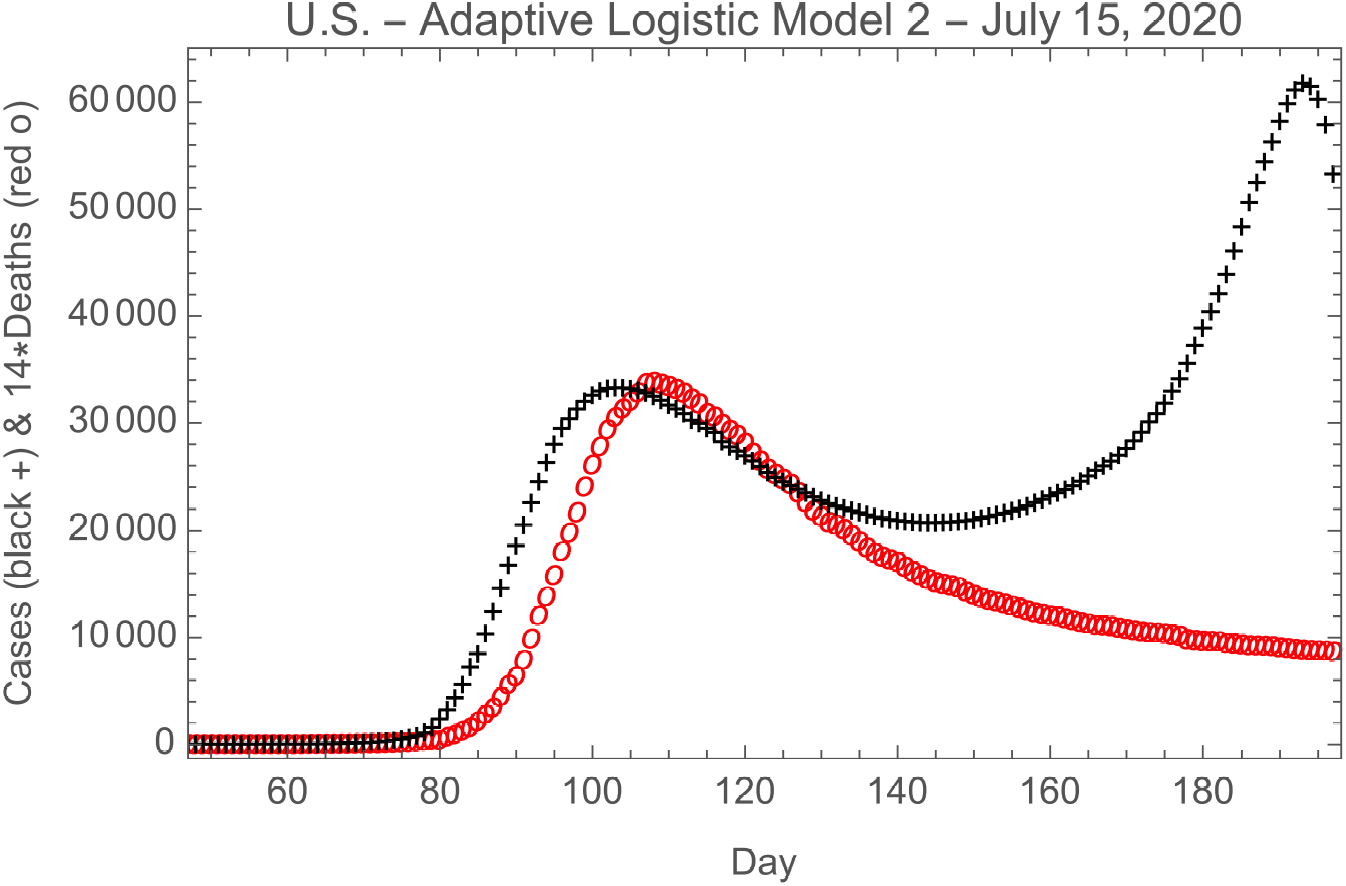
Comparison of the ALM-2 fits to daily cases and daily deaths due to COVID-19 in the U.S. The black pluses are the fit to the cases data and the red circles are 14 times the fit to the deaths data. Day 1 is January 1, 2020.

**Fig. 6.**
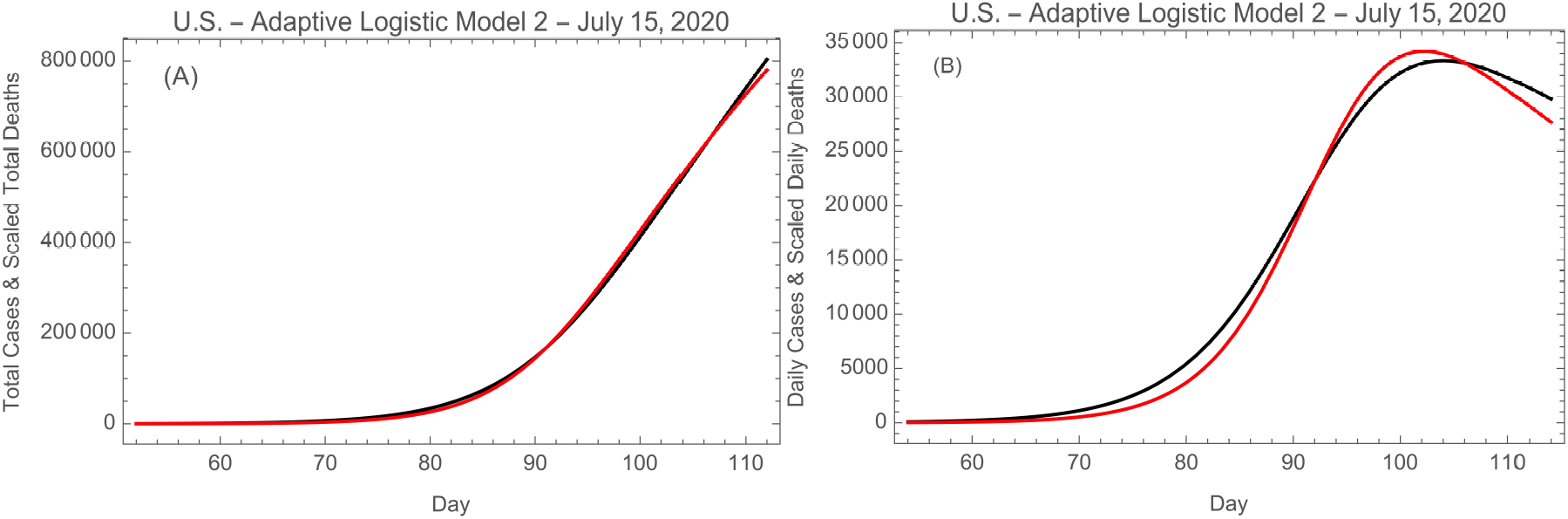
(A) Shifted and scaled total cases and deaths models, and (B) shifted and scaled daily cases and deaths models. In both plots the number of cases is the black curve and the number of deaths is the red curve. Day 1 is January 1, 2020.

Figure 7 shows the evolution of the number of cases and the number of deaths as a function of the effective rate constant *r*_eff_, with day number as the parameter. The curves start in the lower right corner with day 1 and continue through day 197. For both cases and deaths, *r*_eff_ initially decreases with time as the numbers of cases and deaths rise. Each curve reaches a maximum when *r*_eff_ is in the range 0.05–0.08, and then the numbers start to decrease. For the number of deaths this continues to the current day. However, there is an inflection point in the cases curve; this represents the point at which both *r*_eff_ and the number of cases begin to increase again. There are hints in the deaths curve and in Fig. 4B that the number of deaths is about to climb.

**Fig. 7.**
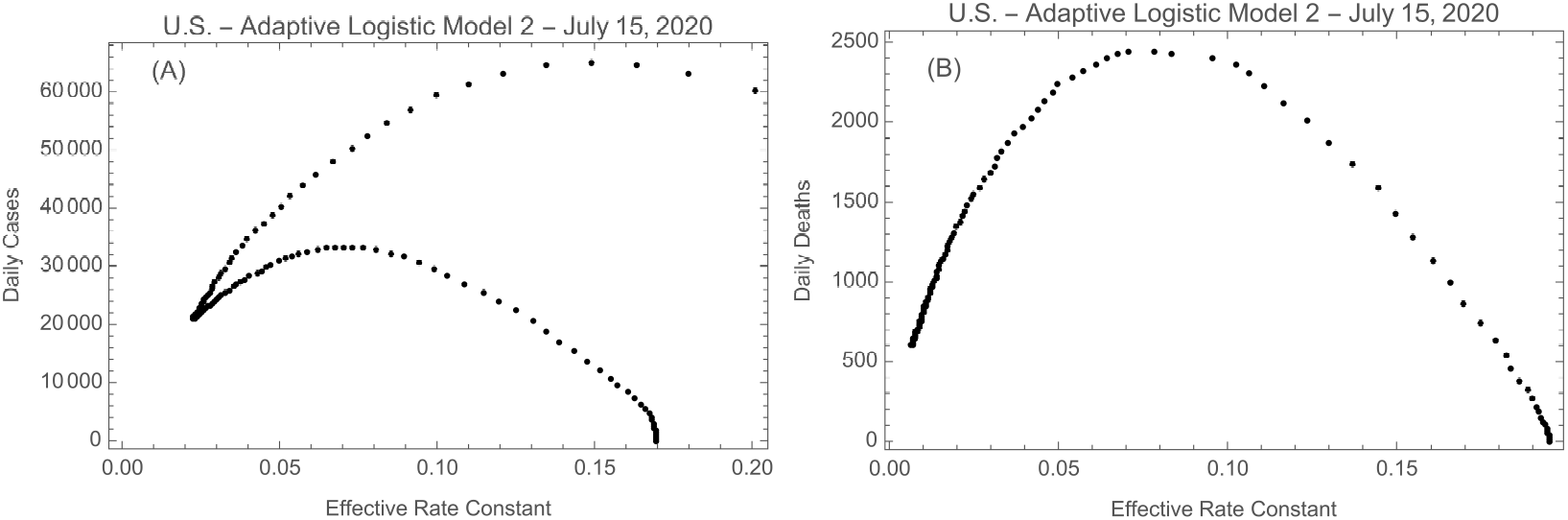
Daily cases (A) and daily deaths (B) as a function of the effective rate constant, showing the evolution in time, starting in the lower right of each plot. The data points are for successive days from 1 to 197.

## 6. Results

The main results of this paper are as follows.

1. The Adaptive Logistic Model can be enhanced by introducing an effective rate constant that decreases at first, due to efforts to mitigate the epidemic, but then begins to increase as these efforts wane. The resulting Adaptive Logistic Model 2 (ALM-2) can account quantitatively for the growth of a second spike of infections in the United States (Fig. 3B).
2. The ALM-2 can account quantitatively for the history of deaths as well, even though no second peak has arisen as yet (Fig. 4).
3. The effective rate constants, as derived from cases and deaths data separately, are shown as functions of time in Fig. 8. From these plots one can see why there is as yet no second peak in deaths–unlike that for the cases, the relevant rate constant for deaths has not reached a minimum and started to rise. Presumably the second peak of deaths has not shown up yet because recently infected individuals, primarily younger people, are less likely to die than were those in the first peak. However, given the new peak of cases we eventually expect a subsequent peak of deaths, and if it occurs the ALM-2 can be applied to that situation.

## 7. Conclusions

It is possible to understand the current (second) peak in COVID-19 cases in the United States in a modified logistic model for epidemics. This new peak is due to relaxation of mitigation factors leading to an increasing effective rate constant *r*_eff_. This can be modeled quantitatively in the Adaptive Logistic Model 2 (ALM-2) using a simple mathematical expression for the initial fall and subsequent rise of *r*_eff_. Unfortunately, this model has limited life because the expression for *r*_eff_ (*g*) cannot be extended to an arbitrarily large number of cases.^4^

**Fig. 8.**
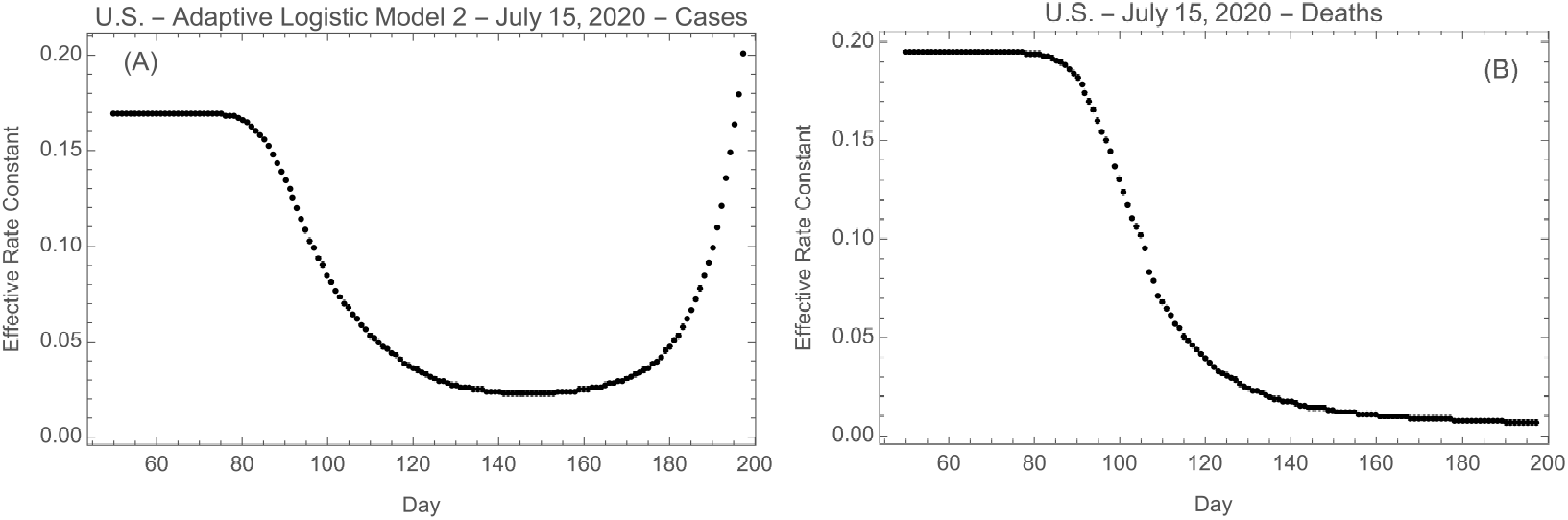
Effective rate constant as a function of time as determined from (A) cases data and (B) deaths data. Day 1 is January 1, 2020.

The same model can be applied to the deaths data; these do not yet show a second peak yet are also successfully described by the ALM-2.

## Data Availability

All of the data come from the European Centre for Prevention and Control of Disease.

https://www.ecdc.europa.eu/en/publications-data/download-todays-data-geographic-distribution-covid-19-cases-worldwide

## 8. Acknowledgements

Brian Boyle, Mary Roberts, and Bob Sauer provided helpful comments and suggestions.

## Footnotes

2 Eq. 3 is so named because it models a population’s adaptation to the disease.

3 This is twice a hyperbolic cosine.

4 A different mathematical form for *r*_eff_, one that asymptotically approaches a constant for large *g*, will have to be used. This was not done here because it involves additional parameters and is thus less well constrained by the current data (Roberts 2020b).

